# Mapping fatigue and its influences on rehabilitation among people with recently acquired spinal cord injury: a scoping review protocol

**DOI:** 10.1101/2023.05.25.23290544

**Authors:** Francesco Giuseppe Materazzi, Paola Paglierani, Laura Simoncini, Ilaria Baroncini, Francesca Serafino

## Abstract

**Background:** Fatigue is a symptom described as a subjective feeling of tiredness, lack of energy and motivation, exhaustion, difficulty in concentrating. Fatigue is a frequent complication among people with spinal cord injury (SCI) and negatively impacts rehabilitative treatment sessions. A multitude of factors contribute to the complexity of fatigue experienced in people with acute and subacute SCI including physical, emotional and cognitive components. Understanding fatigue and fatigability mechanisms may have implications in optimizing perceived exertion during rehabilitation treatment for promoting better functional recovery after SCI. Despite the relevance of this topic, no study has been conducted to map the role of fatigue in acute and subacute SCI. Therefore, the aims of the present scoping review are: to map the currently available evidence describing how fatigue is defined and conceptualized and how to influence the rehabilitation in acute and subacute patients with spinal cord injury. In addition, an overview of the most used assessment tools will be provided.

**Inclusion criteria:** Studies considering people with recently acquired SCI (up to 1 year from the date of the event) and looking at any aspect of fatigue will be eligible for inclusion. Any rehabilitation treatment sessions reported by each study and any context will be considered.

**Methods:** This scoping review will be performed in accordance with the Joanna Briggs Institute guidance. MEDLINE, Cochrane Central, CINAHL Complete and PEDro databases will be searched from inception to April 2023. Additional studies will be identified through searching in gray literature and the reference lists of relevant articles. No study design, time, geographical, setting restrictions will be applied. We will include only studies published in English.

**Conclusions:** This will be the first scoping review to provide a comprehensive overview of all studies dealing with any aspect of fatigue in people with recently acquired spinal cord injury. The results will add meaningful information for clinicians about the management of this symptom and its influences on rehabilitation and for researchers to direct future research. Moreover, an overview of the most used assessment tools to investigate fatigue in SCI will be provided.

## INTRODUCTION

Fatigue is a frequent complication among people with spinal cord injury (SCI) and negatively impacts rehabilitative treatment sessions. It’s the second most frequent limiting factor that occurs during rehabilitation, only preceded by pain [1]. Despite the recognized importance of this problem in people with SCI, there are still considerable knowledge gaps regarding the underlying mechanisms of fatigue[2]. This is, in some measure, a consequence of confusing terminology ranging from subjective feeling of tiredness, lack of energy and motivation, to temporary impairments in neuromuscular function and muscle force production[3]. Furthermore, generally a multitude of factors contribute to the complexity of fatigue experienced in people with acute and subacute SCI including physical, emotional and cognitive components[4]. Based on the recent conceptual framework proposed by Enoka et al.[5,6], fatigue can be conceptualized as a disabling self-reported symptom with two interdependent attributes of performance fatigability and perceived fatigability. Performance and perceived fatigability are both relevant because either can limit physical and cognitive function. Understanding fatigue and fatigability mechanisms may have implications in optimizing perceived exertion during rehabilitation treatment for promoting better functional recovery after SCI.

## METHODS

The present scoping review will be conducted in accordance with the guidance developed by members of the Joanna Briggs Institute (JBI) Scoping Review Methodology Group[7]. The Preferred Reporting Items for Systematic reviews and Meta-Analyses extension for Scoping Reviews (PRISMA-ScR) Checklist for reporting will be used to report the final paper[8]. We will follow the framework of Population, Concept and Context (PCC) proposed by the JBI to describe the elements of the inclusion criteria.

### Review question(s)

We formulated the following research question: “What is known from the existing literature about the evidence regarding fatigue and its influences on rehabilitation among people with recently acquired spinal cord injury?”

The objectives of this scoping review will be to:

1. Provide a comprehensive overview of all studies describing how fatigue is defined and conceptualized in people with recently acquired spinal cord injury;
2. Investigate how fatigue influences rehabilitation in acute and sub-acute patients with spinal cord injury;
3. Identify and analyze the most used assessment tools measuring fatigue utilized in the included studies.

### Inclusion criteria

Studies will be eligible for inclusion if they meet the following PCC criteria.

- Population: Participants with spinal cord injury complete and incomplete (including non-traumatic and traumatic). Up to 1 year from the date of the event. Adult patients (16+ years).
- Concept: Multidimensional nature of fatigue (perceived and performance fatigability).
- Context: Any rehabilitation treatment sessions.

#### Sources

This scoping review will consider any study designs or publication type including primary research studies, reviews, guideline implementation. No time, geographical, setting restrictions will be applied. We will include only studies published in English.

### Exclusion criteria

Studies that do not meet the specific PCC criteria or provide insufficient information will be excluded. In particular, articles with participants suffering at the same time from multiple sclerosis spina bifida, SLA, Parkinson disease, stroke and other neurologic illness will be excluded.

### Search strategy

A preliminary search in MEDLINE and Google scholar was undertaken to identify articles on the topic and used keywords. The information gained from the initial search was used to develop a more comprehensive search strategy based on the PCC framework. We analyzed all the terms regarding fatigue in SCI patients during any rehabilitation treatments. A final comprehensive search was conducted across MEDLINE (through PubMed interface). The search strategy, including all identified keywords and index terms, was adapted for use in Cochrane Central, CINAHL Complete and PEDro (see Table 1, which displays the search strategies). In addition, gray literature (Google scholar) and the reference lists of all relevant studies will be searched.

**Table 1.**
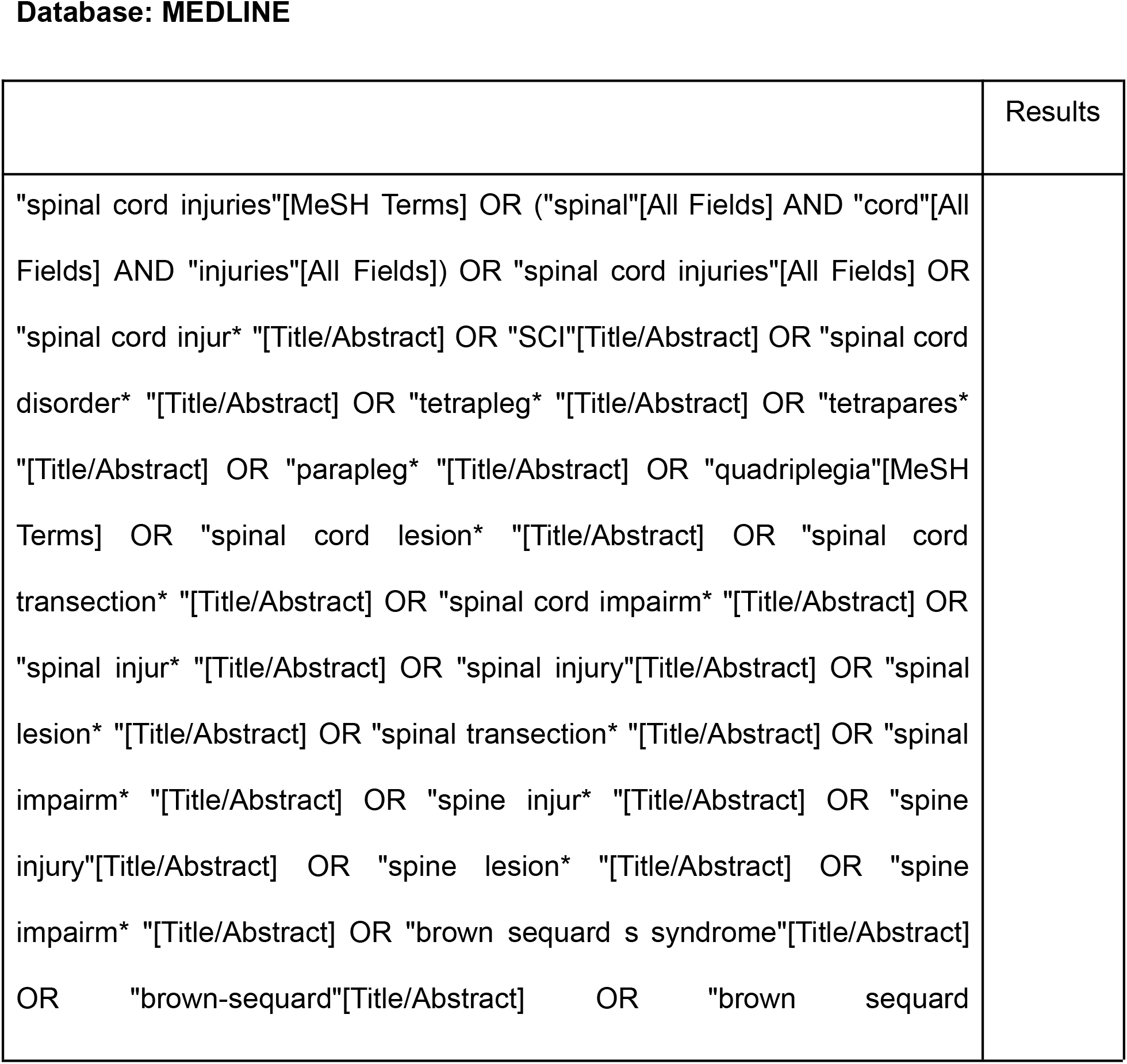
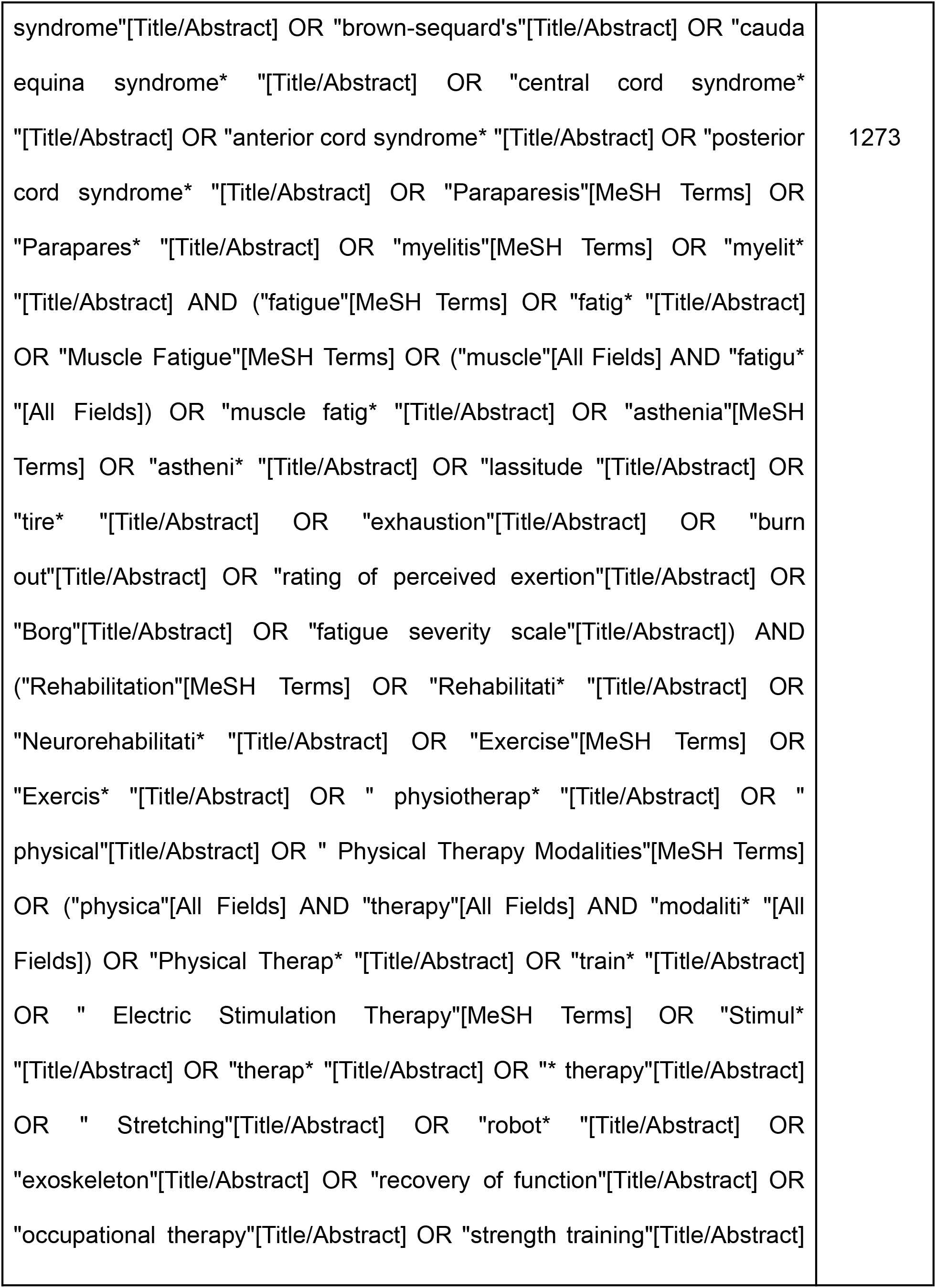
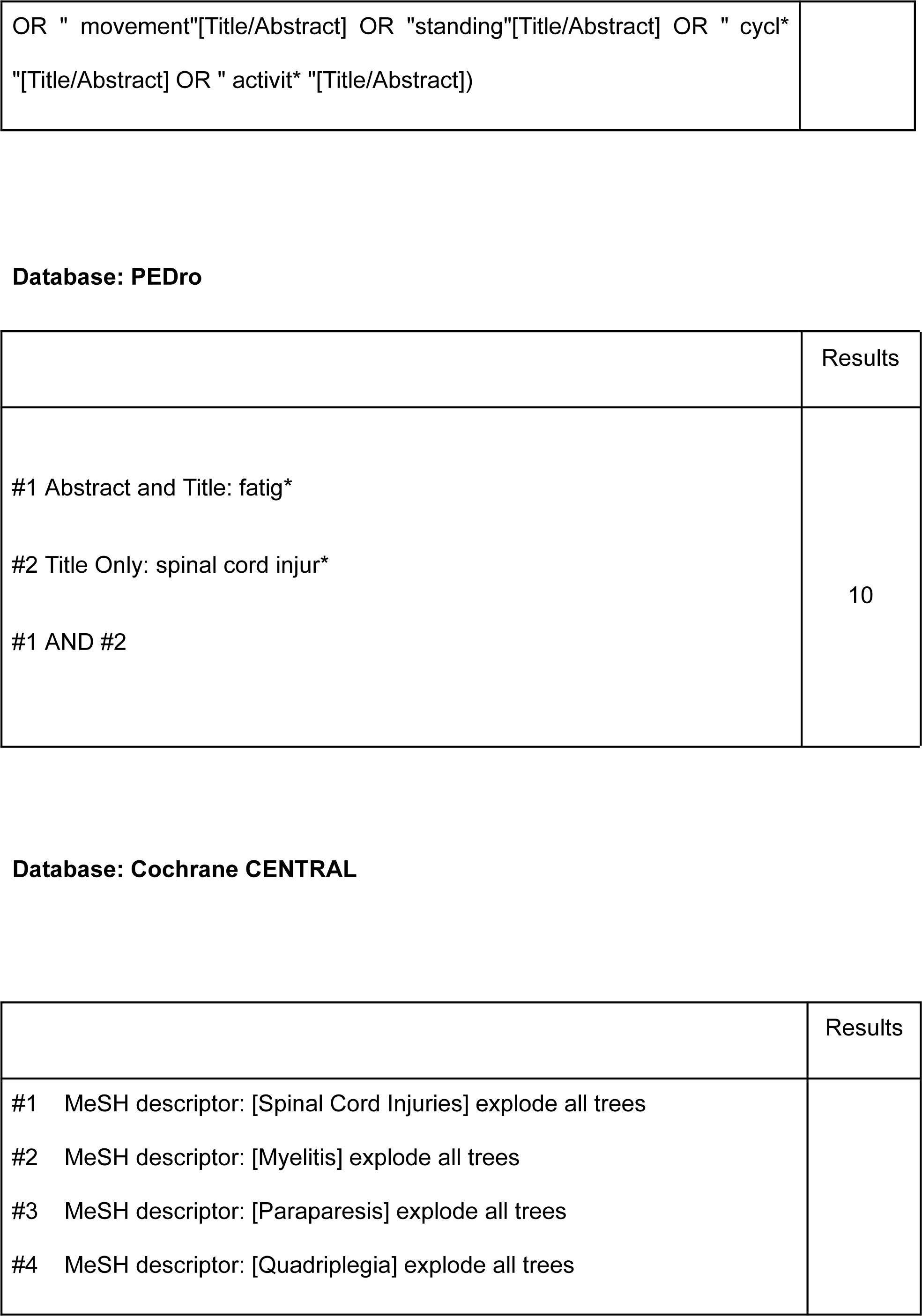
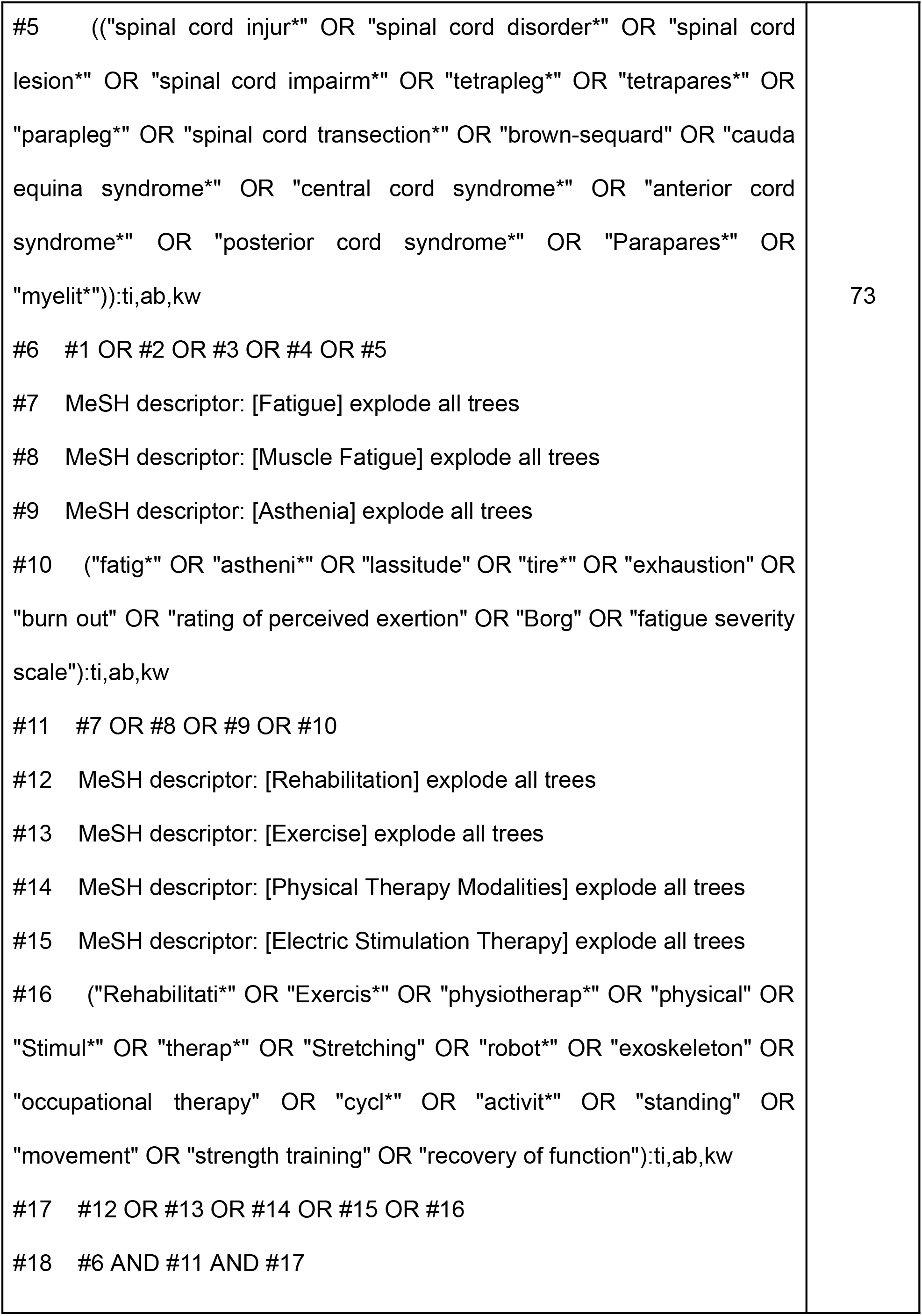
Search strategies of searched databases Topic: The effect of fatigue on rehabilitation in people with recently acquired spinal cord injury Search date: April 7, 2023 No date and language restrictions.

### Study selection

Once the search strategy has been successfully completed, search results will be collated and imported to Zootero. Duplicates will be removed using the software before the file containing a set of unique records is made available to reviewers for further processing (study screening and selection). The review process will consist of two levels of screening using Rayyan QCRI online software[9]: (1) a title and abstract review and (2) a full-text screening. For both levels, two investigators will screen the articles independently to determine if they meet the inclusion/exclusion criteria. In case of any disagreement on inclusion, both reviewers will review full-text articles again. If an agreement cannot be reached, this will be resolved by an independent third reviewer adjudication.

Reasons for the exclusion of any full-text source of evidence will be recorded and reported in the scoping review report. The results of the search will be reported and presented in the latest published version of the Preferred Reporting Items for Systematic Reviews and Meta-analyses (PRISMA) flow diagram (see Figure 1).

**Figura 1.**
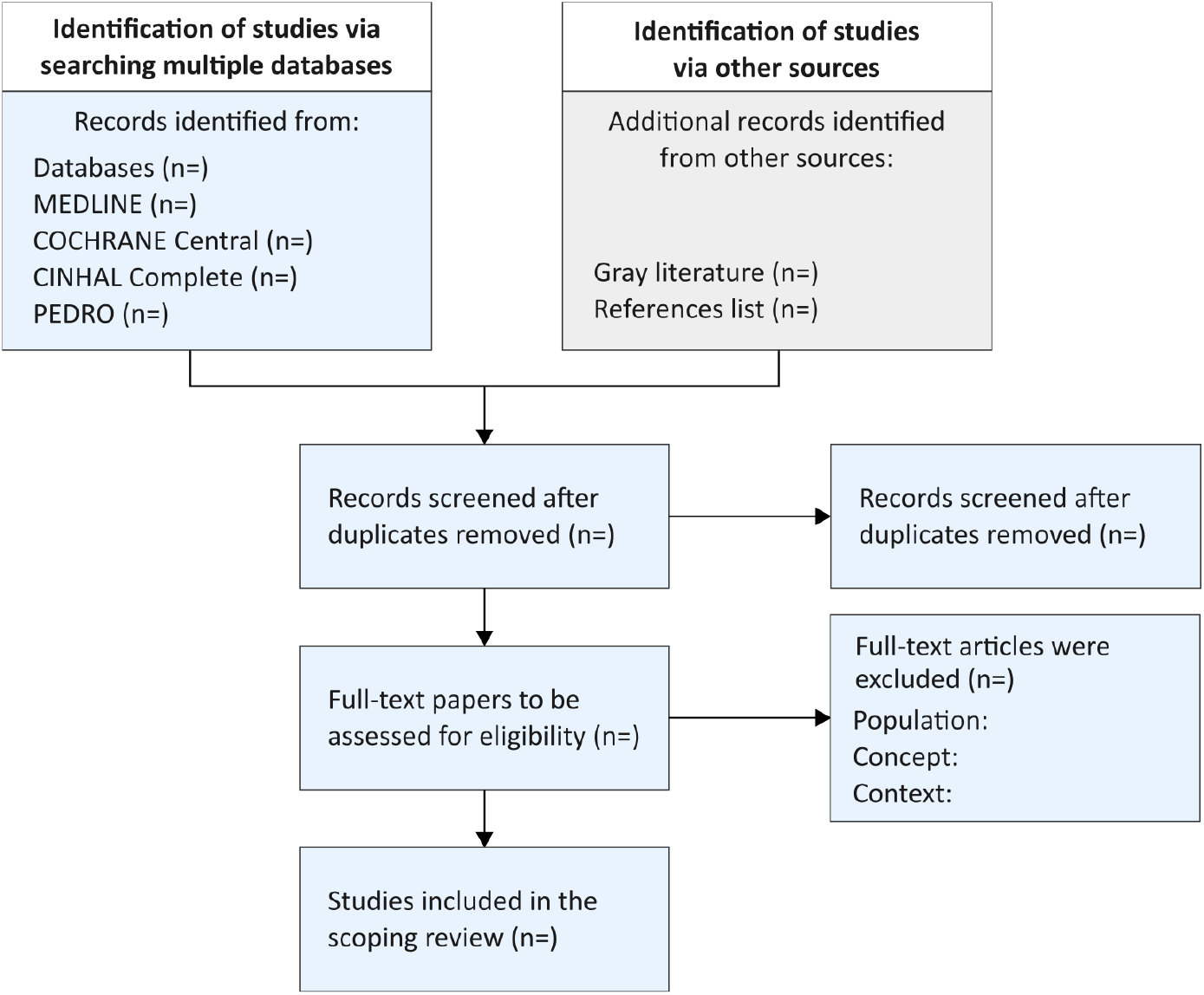
Flow diagram

### Data extraction

An ad-hoc data collection form will be developed by the reviewers and key information (e.g. title, authors, country, year of publication, type of study, concept of fatigue used, measurement tools) on the included studies will be collected. A draft collection tool is provided (see Table 2, which illustrates the data extraction instrument).

**Table 2.** Data extraction instrument: draft

|  |
| --- |
| Study details |
| Title |
| Author(s) |
| Year of publication |
| Study design |
| • If correlation study → factors in relation with fatigue<br>• If intervention study → intervention type and comparator; duration of the intervention |
| Aim(s) of the study |
| Population |
| Sample size |
| Age |
| Characterization of spinal cord injury<br>(ASIA, complete/incomplete, traumatic/non traumatic) |
| Time since injury (months) |
| Concept |
| “Fatigue” definition described by authors |
| Conceptual information about fatigue |
| Measurement tool(s) used relative fatigue |
| Outcome measures |
| Context |
| Country |
| Setting |
| Results/ main key point(s) |

This form will be reviewed by the research team and pre-tested by all reviewers before implementation to ensure that the form captures the information accurately. Charting results is commonly an iterative process during scoping reviews; other data can be added to this form according to the subgroups that could emerge from the analysis of the studies included. Modifications will be detailed in the full scoping review.

### Data synthesis

The results will be presented numerically and thematically. Studies identified and included will be reported, and the description of the search decision process will be mapped. In addition, extracted data will be summarized in tabular and diagrammatic form. A thematic summary will be performed pertaining to themes and key concepts relevant to the research questions.

## Data Availability

All data are included in the report

## Data Availability

All data are included in the report.

## List of abbreviations

SCI: Spinal cord injury
JBI: Joanna Briggs Institute
PCC: Population-concept-context
PRISMA-ScR: The Preferred Reporting Items for Systematic reviews and Meta-Analyses extension for Scoping Reviews

